# Geospatial Patterns of Progress towards UNAIDS “95-95-95” Targets and Community Vulnerability in Zambia

**DOI:** 10.1101/2023.04.24.23289044

**Authors:** Diego F Cuadros, Tuhin Chowdhury, Masabho Milali, Daniel Citron, Sulani Nyimbili, Natalie Vlahakis, Theodora Savory, Lloyd Mulenga, Suilanji Sivile, Khozya Zyambo, Anna Bershteyn

## Abstract

In sub-Saharan Africa, HIV/AIDS remains a leading cause of death. The UNAIDS established the “95-95-95” targets to improve HIV care continuum outcomes. Using geospatial data from the Zambia Population-based HIV Impact Assessment (ZAMPHIA), this study aims to investigate geospatial patterns in the “95-95-95” indicators and individual-level determinants that impede HIV care continuum in vulnerable communities, providing insights into the factors associated with gaps. This study used data from the 2016 ZAMPHIA to investigate the geospatial distribution and individual-level determinants of engagement across the HIV care continuum in Zambia. Gaussian kernel interpolation and optimized hotspot analysis were used to identify geospatial patterns in the HIV care continuum, while geospatial k-means clustering was used to partition areas into clusters. The study also assessed healthcare availability, access, and social determinants of healthcare utilization. Multiple logistic regression models were used to examine the association between selected sociodemographic and behavioral covariates and the three main outcomes of study. Varied progress towards the “95-95-95” targets were observed in different regions of Zambia. Each “95” displayed a unique geographic pattern, independent of HIV prevalence, resulting in four distinct geographic clusters. Factors associated with gaps in the “95s” include younger age, male sex, and low wealth, with younger individuals having higher odds of not being on ART and having detectable viral loads. Our study revealed significant spatial heterogeneity in the HIV care continuum in Zambia, with different regions exhibiting unique geographic patterns and levels of performance in the “95-95-95” targets, highlighting the need for geospatial tailored interventions to address the specific needs of different subnational regions. These findings underscore the importance of addressing differential regional gaps in HIV diagnosis, enhancing community-level factors, and developing innovative strategies to improve local HIV care continuum outcomes.

## 1 Introduction

Maintaining undetectable HIV viral load via life-long antiretroviral therapy (ART) is recommended for all people living with HIV (PLHIV) as it has been shown to preserve health and reduce onward transmission.[1] However, despite these recommendations being adopted into clinical guidelines, HIV/AIDS remains a leading cause of death in sub-Saharan Africa (SSA), where two-thirds of the world’s PLHIV reside.[2] This is due to substantial numbers of PLHIV remaining undiagnosed, being diagnosed but not receiving ART, or receiving ART but not achieving viral load suppression.[3] In response, UNAIDS established operational targets for national HIV programs to diagnose 95% of those living with HIV, provide ART to 95% of those diagnosed, and achieve viral load suppression in 95% of those receiving ART, known as the “95-95-95” targets.[4] To assess progress towards these targets, Population-based HIV Impact Assessments (PHIAs) have been conducted, providing nationally representative cross-sectional data on HIV diagnosis, treatment, and viral load suppression, as well as demographic and health data.[5] However, questions remain about the uniformity of progress across countries and the extent to which “95-95-95” indicators correlate with one another and with the epidemiology of the HIV/AIDS pandemic.

The HIV epidemic in Zambia is one of the most severe in the world, with an estimated 1.3 million people living with HIV in 2020, according to UNAIDS.[6] Despite progress in the HIV response, with a decline in new infections from 58,000 in 2010 to 38,000 in 2020, Zambia still faces significant challenges in achieving the “95-95-95” targets.[5] Using geospatial data from the PHIA in Zambia, a country with a high HIV burden, we investigated the geospatial patterns in “95-95-95” indicators and their relationship to HIV prevalence and individual level determinants that could impede HIV diagnosis, treatment, and viral load suppression in these vulnerable communities.

The main objective of this study is to provide insights into the extent to which progress towards “95-95-95” has been even or uneven across Zambia and identify regions that require differential emphasis on HIV diagnosis, treatment, or viral load suppression to accelerate progress. Using geospatial and statistical analyses of individual-level determinants of engagement across the “95-95-95” continuum, this study aims to provide insights into the factors associated with gaps in the HIV care continuum not only in Zambia but also in other sub-Saharan African countries. The study was conducted in collaboration with the Zambia Ministry of Health, highlighting the importance of partnerships between governments and researchers in addressing the HIV epidemic in SSA.

## 2 Methods

### 2.1 Data sources

This analysis uses individual-level questionnaires and biomarker data on adults ages 15–59 participating in the 2016 Zambia Population-based HIV Impact Assessment (ZAMPHIA), which is the most recent ZAMPHIA dataset publicly available.[7] ZAMPHIA is a nationally representative cross-sectional survey of households across all 10 provinces of Zambia, as well as geospatial data on communities included in the survey. ZAMPHIA used a two-stage stratified sample design, first selecting 511 enumeration areas (EA) from the 2010 Census of Population and Housing in Zambia, and then inviting households in each EA to participate in the survey.[5, 8] Households consenting to participate in the survey received face-to-face structured household and individual interviews, home-based HIV testing with immediate return of results and further laboratory testing, and viral load testing. HIV testing used the Alere Determine™ HIV-1/2 rapid test (Abbott Laboratories, Lake Bluff, IL) and positive results were confirmed using the Uni-Gold™ HIV-1/2 rapid test (Trinity Biotech, Wicklow, Ireland). Positive specimens were also tested for the presence of the antiretroviral drugs (ARVs) efavirenz, nevirapine, lopinavir, and atazanavir, as described elsewhere.[7] The latitude and longitude of the approximate centroid of each EA was recording using the global positioning system (GPS) (Supplementary Figure 1). Anonymized data from ZAMPHIA are available to researchers with approval from the Zambia Ministry of Health, US Centers for Disease Control, and the International Center for AIDS Care and Treatment Program (ICAP) at Columbia University. Survey questionnaires, consent forms, and aggregated results are publicly available.[7]

### 2.2 Main outcomes

#### 2.2.1 HIV status

Positive HIV status was defined as positive results on initial and confirmatory HIV tests. Individuals with negative or discrepant HIV test results were not classified as HIV-positive.

#### 2.2.2 Awareness of positive HIV status

For participants classified as HIV-positive, awareness of HIV status was assigned based on multiple survey indicators. Participants were assumed to be aware of their HIV positive status if any of the following were true: (1) they reported their HIV status as positive on the survey questionnaire, (2) if ARVs were detectable in their blood sample, or (3) HIV viral load was <1000 HIV RNA copies per milliliter in their blood sample.

#### 2.2.3 ART status

For participants classified as aware of their positive status, ART status was assigned based on multiple survey indicators. Participants were assumed to be receiving ART if (1) they report to receive ART (2) one or more of the tested ARVs was detected, or (3) HIV viral load was <1000 HIV RNA copies per milliliter.

#### 2.2.4 Viral load suppression

For participants classified as receiving ART, viral load suppression was defined as having <1000 HIV RNA copies per milliliter in their blood sample.

### 2.3 Spatial analysis

#### 2.3.1 Mapping of the care continuum

Spatial analysis combined the prevalence individual-level care continuum variables with EA-level GPS coordinates. We performed Gaussian kernel interpolation to create spatially smoothed estimates of HIV prevalence, HIV status awareness prevalence among those HIV-positive, ART prevalence among those aware of status, and viral load suppression prevalence among those receiving ART. Estimates were aggregated at the district for each of Zambia’s 116 administrative districts to increase relevance to health policy decision-making. We performed optimized hotspot analysis [9, 10] to identify the geospatial structure of the prevalence of HIV along with the prevalence of the three UNAIDS targets, with the identification of hotspots (areas with high HIV prevalence and high percentages of HIV awareness, ART uptake, and viral load suppression) and coldspots (areas with low HIV prevalence and low percentages of HIV awareness, ART uptake, and viral load suppression).

#### 2.3.2 Geospatial visualization of care continuum clustering

Multivariable spatial visualization of the care continuum, HIV prevalence, HIV status awareness, ART status, and viral load suppression, were ascertained using a geospatial k-means clustering approach, partitioned districts so that within-cluster similarity is maximized and between-cluster similarity minimized (or dissimilarity maximized). We categorized districts into k=4 clusters using the open-source spatial data science tool GeoDa, version 1.20 environment,[11] as described elsewhere.[12-14] For each cluster, we reported the mean of each care continuum variable. Interquartile ranges (IQRs) represent the variation across the districts included in the cluster. Maps of care continuum metrics by district and cluster were generated using ArcGIS Pro version 3.1.[15]

#### 2.3.3 Mapping of healthcare availability, access, and social determinants of healthcare utilization

For each cluster, we additionally assessed levels of healthcare availability and access using data from the Africa COVID Community Vulnerability Index (https://precisionforcovid.org/africa). [16] The index combines several factors associated with healthcare availability (health system strength, health system capacity, and access to health care) and community vulnerability and access to healthcare (household crowding, improved housing, sanitation, access to transportation, and road connectivity), as well as socioeconomic factors that can act as determinants of healthcare utilization (access to information, education, poverty, and unemployment). Maps of the results were generated using ArcGIS Pro Version 3.1.[15]

#### 2.3.4 Statistical analysis

We assessed the association at individual level between selected sociodemographic and behavioral covariates and the three main outcomes of study by fitting three different models for HIV status awareness, ART status, and viral load suppression. Behavioral and sociodemographic covariates included gender, age, wealth index, religion (traditional, catholic, other), age at the first sexual contact, marital status, place of residence (urban or rural), work in the last 12 months, and away from home in the last 12 months. Odds ratio was calculated by exponentiating the coefficient of multiple logistic regression.

We conducted the individual-level analysis separately using each of the three HIV care continuum outcomes as dependent variables. For Model 1: HIV status awareness, the survey data was subsampled to include HIV-positive individuals only; For Model 2: ART status, the survey data was subsampled to include HIV-positive individuals aware of their status only; and for Model 3: viral load suppression the survey data was subsampled to include HIV-positive individuals aware of their status and on ART treatment. Population subsets for each model were generated using the ‘subset’ argument to the svygl() function in the R package survey. We fitted survey-weighted multiple logistic regression models with the svyglm() function to account for the multistage sampling survey design of ZAMPHIA.[17]

#### 2.3.5 Ethical considerations

This analysis uses available de-identified data from the ZAMPHIA, which was funded by PEPFAR with technical assistance through the US Centers for Disease Control and Prevention (CDC) under the terms of the cooperative agreement U2GGH001226. Human subjects and ethical approval for the ZAMPHIA survey was granted by the Zambia National Health Research Ethics Board, the Tropical Diseases Research Centers Ethics Review Committee, and the Institutional Review Boards at the Centers for Disease Control and Prevention (CDC; Atlanta, Georgia, USA). All participants provided written informed consent or assent for the original survey.

#### 2.3.6 Data availability statement

Data are available in a public, open-access repository. The data that support the findings of this study are available from the Population-Based HIV Impact Assessment (PHIA; https://phia-data.icap.columbia.edu/), but restrictions apply to the availability of these data, which were used under license for the current study and so are not publicly available. We sought and were granted permission to use the core data set for this analysis by PHIA.

#### 2.3.7 Funding

Research reported in this publication was supported by the National Institute of Mental Health and the National Institute of Allergy and Infectious Diseases of the National Institutes of Health under award numbers 5R01MH124478 and 1R01AI174932. The content is solely the responsibility of the authors and does not necessarily represent the official views of the National Institutes of Health.

## 3 RESULTS

### 3.0.1 National-level HIV care continuum

Nationally, HIV prevalence estimated among adults ages 15–59 in Zambia was 12.3% (95% CI: 11.8– 12.8). HIV prevalence among female adults was 14.9% (95% CI: 14.3–15.7), and among male adults was 9.4% (95% CI: 8.9–9.9). HIV prevalence among adults in rural areas was 15.7% (95% CI: 14.9–16.5) and in urban areas was 9.5% (95% CI: 8.9–10.1). HIV prevalence was highest in the 45–54 age group at 23.9% (95% CI: 21.7–26.2) and lowest in the 15–24 age group at 3.9% (95% CI: 3.4–4.3). Nationally, 72.3% (95% CI: 70.3–74.2) of HIV-positive adults were aware of their status, 87.3% (95% CI: 85.5– 89.1) of those aware of their status were receiving ART, and 89.5% (95% CI: 87.7–91.2) of those receiving ART had suppressed viral load.

### 3.0.2 Geospatial patterns of the HIV care continuum

Geospatial patterns of the HIV prevalence and care continuum engagement differed markedly across stages of the HIV care continuum (Figures 1 and 2). HIV prevalence (Figure 1A and 2A) was highest in the central axis of the country, spanning the Copperbelt and Central provinces and parts of the Central and Western provinces, with an identified hotspot where prevalence exceeded 14% of all adults. HIV prevalence was lowest in the north-western and eastern parts of the country, especially Muchinga province.

**Figure 1.**
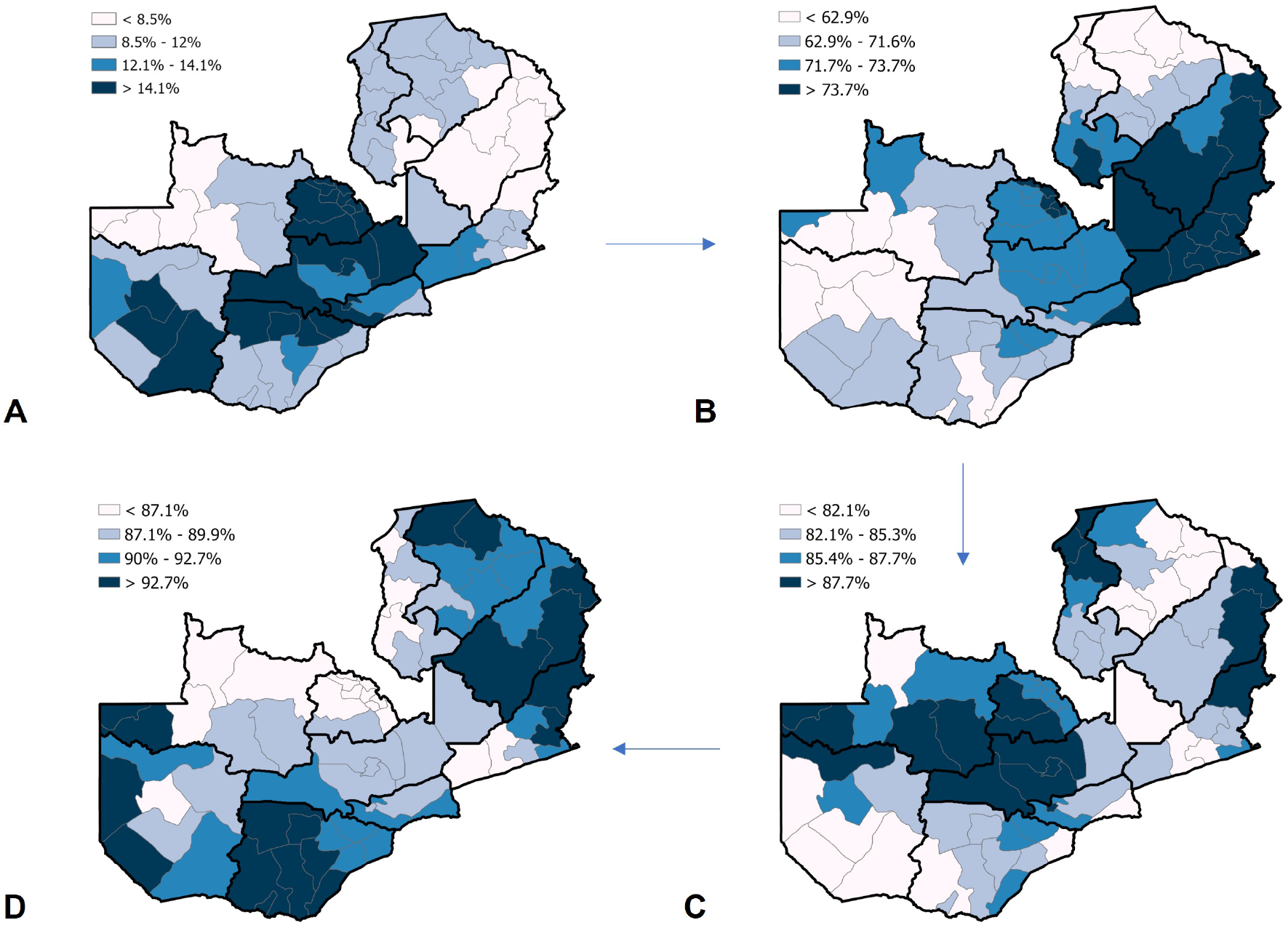
Prevalence distributions of A) HIV prevalence; B) Prevalence of HIV positive individuals aware of their status; C) prevalence of HIV positive individuals aware of their status on ART treatment; and D) prevalence of HIV positive individuals aware of their status on ART treatment that are viral load suppressed

**Figure 2.**
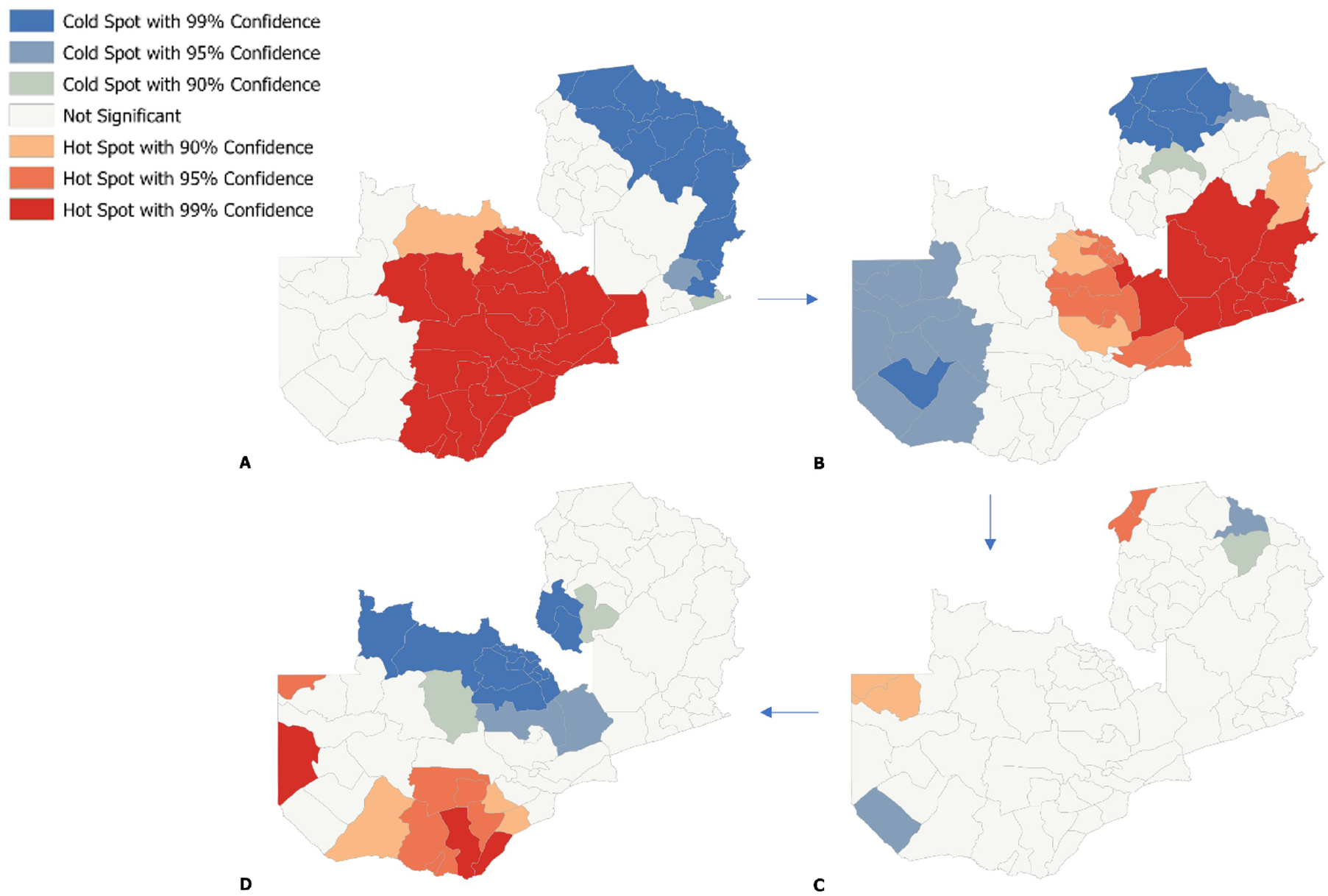
Hotspot maps of A) HIV prevalence; B) Prevalence of HIV positive individuals aware of their status; C) prevalence of HIV positive individuals aware of their status on ART treatment; and D) prevalence of HIV positive individuals aware of their status on ART treatment that are viral load suppressed

Awareness of positive HIV status among those living with HIV (Figures 1B and 2B) was the largest gap in the HIV care continuum across geographies. It exhibited a markedly different geospatial pattern than the pattern of HIV prevalence, with a gradient from low status awareness in the west to high status awareness in the east. Awareness of status was lowest in the western half and north-eastern quadrant of the country, and highest in the southeastern quadrant. In the central axis of the country where HIV prevalence is concentrated, awareness of status was at an intermediate level relative to elsewhere in the country.

ART coverage and viral load suppression (Figures 1C and 1D) also exhibited unique geospatial patterns. ART coverage among those aware of their status was highest in a band from the center to the west of the country and in portions of its northeastern and eastern borders and was lowest in the south and portions of the east (Figure 1C). Viral load suppression among those receiving ART clustered in the east, west, and south, and lowest in the north and center of the country (Figures 1D and 2D).

Taken together, these geographic patterns of HIV prevalence, status awareness, ART status, and viral load suppression could be clustered into four regions with distinct patterns (Figure 3). Cluster 1¬ had the highest HIV prevalence (mean: 14.1%, IQR: 11.2-16.4) and had a relatively high proportions of status-aware PLHIV on ART (mean: 85.4%, IQR: 83.2-87.7) and viral load suppression among those on ART (mean: 86.7%, IQR: 85.7-88.6).

**Figure 3.**
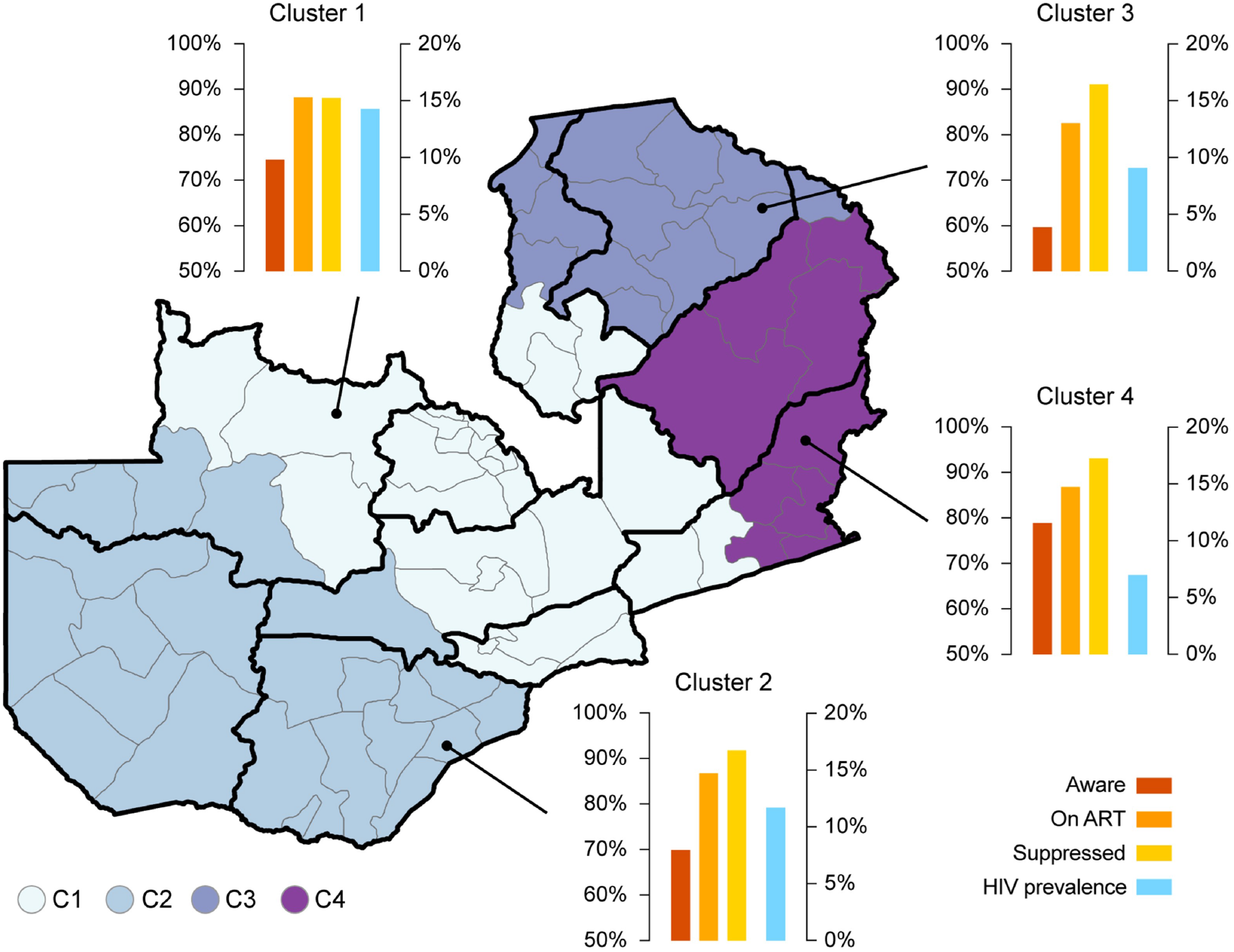
Spatial structure of the HIV epidemic and HIV care continuum in Zambia

Cluster 2 includes districts with a range of HIV prevalence (mean: 12.1%, IQR: 10.2-14.5) and has low HIV status awareness (mean: 63.7%, IQR: 58.1-68.7), low ART coverage among those aware of status (mean: 84.2%, IQR: 81.7–86.9), and relatively high viral load suppression (mean: 93.14%, IQR: 91.1–95.3). Community vulnerability index was high in Cluster 2 compared to the national average (Supplementary Figure 3), with many districts in the top quartile for all three key indicators of health systems (Supplementary Figure 3A), transportation and housing (Supplementary Figure 3B), and socioeconomic factors (Supplementary Figure 3C).

Cluster 3 had moderate HIV prevalence (mean: 9.2%, IQR: 8.5–10.2) and the lowest HIV status awareness (mean: 57.7%, IQR: 48.2–66.7) and ART coverage (mean: 83.4%, IQR: 78.8–87.6) of any cluster. Like Cluster 2, this cluster had a high average community vulnerability index, with nearly all of Cluster 2 in the top quartile of health systems vulnerability (Supplementary Figure 3A), and some sub-regions in the quartile for vulnerability in terms of transportation and housing (Supplementary Figure 3C) or socioeconomic factors (Supplementary Figure 3A). The high community vulnerability in terms of health systems could explain low rates of HIV diagnosis, and low ART coverage among those diagnosed.

Finally, Cluster 4 had the lowest HIV prevalence (mean: 6.9%, IQR: 5.3–8.4) and the highest HIV status awareness (mean: 81.1%, IQR: 74.4–87.1) and viral load suppression (mean: 92.3%, IQR: 92.5–93.3) of any cluster. Much of this cluster was also in the top quartile for community vulnerability in terms of health systems (Supplementary Figure 3A) and transportation and housing (Supplementary Figure 3B), despite relatively strong performance along the HIV care continuum.

### 3.0.3 Factors associated with HIV care continuum status

To explore why each of the three “95s” exhibits such different geospatial patterns, we conducted statistical analyses for assess individual-level predictors of gaps in each “95,” (Figure 4) outside of the geographic sectors explored in the previous analysis. Overall, we found that gaps in all three “95s” were associated with younger age, male sex, and low wealth (though sex and wealth were not statistically significant for ART use among those diagnosed). Marital status and residence were only associated with diagnosis, while travel away from home was only associated with viral load suppression among those on ART. None of the “95s” were significantly associated with education, religion, employment, or age at first sex. For gaps in the first “95,” we found that gender, age, wealth index, marital status, and place of residence were significantly associated with the odds of HIV status awareness. Males had higher odds of been unaware of the HIV status compared to females, and odds of being unaware of status was highest in the youngest age group analyzed (15–24) and declined with age. Odds of being unaware of positive HIV status were highest in the lowest wealth quintile, in individuals who were divorced/separated/widowed or never married, and those with rural residences. For gaps in the “second 95,” we found that only age was associated with not on ART. Younger PLHIV individuals aware of their status had higher odds of not been on ART. Other factors that were significantly associated with lack of diagnosis were not significantly associated with lack of ART after diagnosis. For gaps in the “third 95,” we found that gender, age, wealth index, and travel away from home were significantly associated with detectable viral load. Odds of having detectable viral load while on treatment were highest for men, younger individuals (ages 15–24 – the youngest age group included in this analysis), those in the lowest wealth quintile, and those who had traveled away from for at least one month during the past 12 months. Fewer than 50% of HIV positive males aged 15–34 and females aged 15–24 known their HIV status. Likewise, less than 65% of HIV positive on ART treatment males aged 15–24 and less than 80% females with the same age had their viral load suppressed (Supplementary Figure 2).

**Figure 4.**
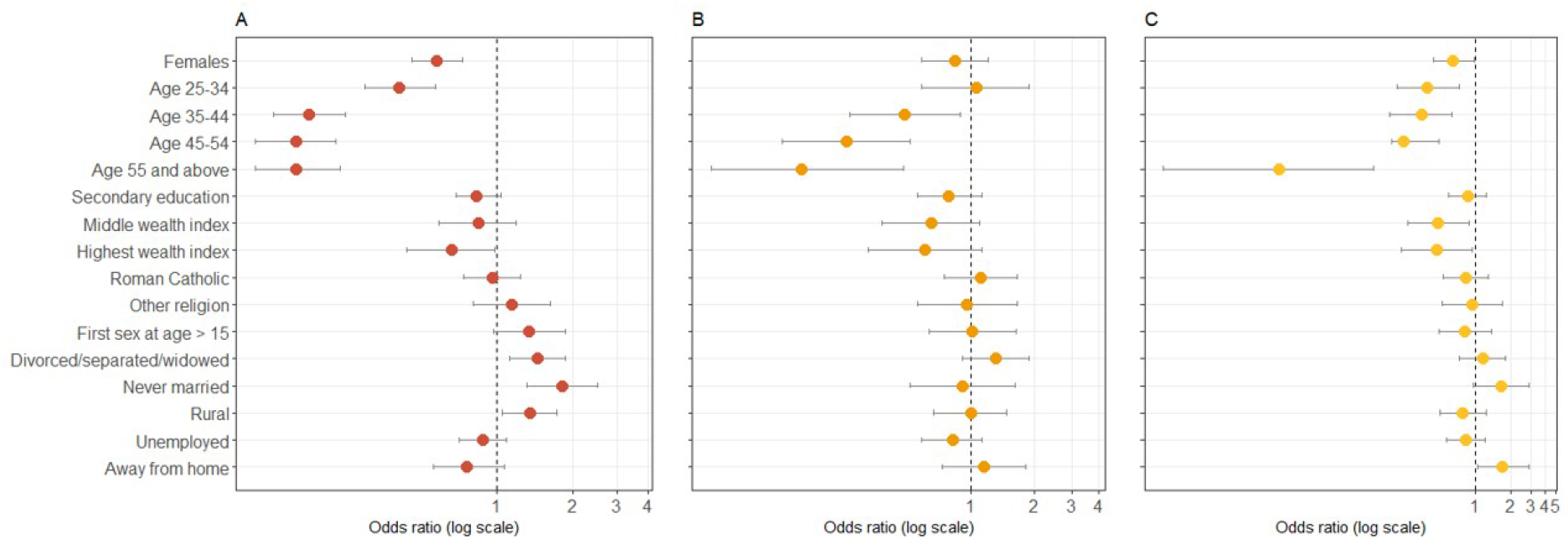
Results from the multivariable regressions for the three different models, (A) Model 1: HIV status awareness, the survey data was subsampled to include HIV-positive individuals only; (B) For Model 2: ART status, the survey data was subsampled to include HIV-positive individuals aware of their status only; and (C) for Model 3: viral load suppression the survey data was subsampled to include HIV-positive individuals aware of their status and on ART treatment

## 4 DISCUSSION

Our study provides valuable insights into the current state of the HIV epidemic in Zambia, highlighting key determinants and geographical disparities of HIV status awareness, ART status, and viral load suppression among HIV-positive individuals. Among adults living with HIV in Zambia, 72.3% had knowledge of their HIV status, with 87.3% of those aware of their HIV status receiving ART treatment, and 89.5% of those receiving ART treatment achieving viral load suppression. However, significant disparities in these metrics were observed across different population groups and geographical regions. Consistent with previous research, our spatial analysis identified pockets of high HIV prevalence in certain regions, such as the Copperbelt and Lusaka provinces.[18–21] Our study also revealed four distinct clusters with unique geographic patterns, exhibiting different levels of performance in the “95–95–95” targets. These clusters included regions with high HIV prevalence, relatively high diagnosis rates, and moderate ART and viral load suppression along the central axis of the country; regions with moderate HIV prevalence, lower diagnosis rates in the south and west, and moderate ART and viral load suppression rates; regions with moderate HIV prevalence, very low rates of diagnosis in the northeast, and low ART and viral load suppression rates; and regions with low HIV prevalence, high performance across all three “95s” in the east. Specifically, Cluster 1, located in the Copperbelt, Central, and Lusaka provinces, has high HIV prevalence rates, relatively high performance in all “95s”, and better healthcare access and infrastructure compared to other regions.[22, 23] Cluster 2, located in the Western and Southern provinces, has moderate HIV prevalence rates but lower rates of diagnosis, ART uptake, and viral suppression than Cluster 1, with limited healthcare access and infrastructure, especially in the Western province.[24-26] Cluster 3, located in the Luapula and Northern provinces, has moderate HIV prevalence rates but very low rates of diagnosis, ART uptake, and viral load suppression, with similar limited healthcare access and infrastructure to Cluster 2.[27] Finally, Cluster 4, located in the Muchinga and Eastern provinces, has the lowest HIV prevalence rates in the country but relatively high performance in all “95s”, especially in the Eastern province, which has relatively good healthcare access and infrastructure.[21, 28] These results underscore the need for tailored interventions to address the specific needs of different subnational regions. Regions with high HIV prevalence may require more resources for HIV testing and diagnosis, while innovative approaches to ART delivery may be necessary in areas with limited healthcare access and infrastructure.

Our study suggests that reporting “95–95–95” targets at the national level may not be sufficient to address subnational care continuum gaps. As noted previously, the largest gaps in each “95” occurred in different geographic regions. Progress towards these targets has been uneven across the country, and gaps vary for each of the “95s”. These results highlight the challenges of achieving the “95-95-95” targets in Zambia, with diagnosing HIV being the most significant obstacle. Therefore, it may be necessary to adopt a tailored approach to healthcare strengthening in different parts of Zambia to improve progress towards HIV treatment targets. We observed that HIV care continuum gaps are not solely related to HIV prevalence, as regions with high HIV prevalence did not necessarily perform better than others, indicating the need for tailored interventions to address the specific needs of different subnational regions in the country to achieve the “95-95-95” targets.

Enhancing community-level factors, such as transportation and housing, may also be critical in improving HIV care continuum outcomes in vulnerable populations. In addition, our study found that care continuum gaps coincided with regions experiencing high community vulnerability, particularly in the domains of healthcare and housing/transportation, indicating that community-level factors are among the drivers of HIV care engagement. Further spatial assessments of community vulnerability could refine our understanding of the contributors to care continuum gaps.

Individual-level factors associated with gaps in the care continuum varied depending on the specific “95” target. Lack of HIV diagnosis was more likely among individuals residing in rural areas and those who were unmarried, while lack of viral load suppression was associated with travel away from home. However, several factors were consistently associated with gaps across all three “95s,” including poverty, male gender, and young age. Females had lower odds of being unaware of their HIV status compared to males, and the odds of being unaware of HIV status decreased with age. Individuals in the highest wealth quintile had lower odds of being unaware of their HIV status than those in the lowest quintile. Age was the only factor associated with not being on ART treatment, and older individuals aware of their HIV status had lower odds of not being on ART treatment compared to those aged 15–24. Similarly, older individuals had lower odds of not being viral load suppressed compared to those aged 15–24 on ART. Individuals in the lowest wealth quintile had higher odds of not being viral load suppressed, and those on ART treatment who were away from home for at least one month during the past 12 months had higher odds of not being viral load suppressed. A comprehensive approach is needed to address gaps in the care continuum. Targeted interventions may be required to increase HIV status awareness and ART uptake among males, rural populations, and individuals from lower wealth quintiles. Additionally, innovative service delivery models, such as mobile ART clinics, may be necessary to improve viral load suppression among individuals who travel away from home, such as providing mobile ART clinics or other innovative service delivery models.

Our study is subject to several important limitations. First, our findings are based on the 2016 ZAMPHIA survey, which represents the most current data publicly available with population-based estimates of “95-95-95”. As a second PHIA study has recently been conducted in Zambia, our research should be repeated once results are available to provide updated estimates. Second, our analysis was limited to the district level, and sub-district level geographic differences may exist. While the PHIA survey provided estimates for certain geographic sub-groups and administrative regions, it was not designed to generate small-area estimates. Future research could improve estimates by incorporating routine health systems data that are larger and more timely, though potentially more biased and incomplete. Finally, our assessment of community vulnerability factors was only qualitative and at a coarse spatial scale. More detailed spatial estimates of community vulnerability could better explain the distinct geographic patterns observed for each “95” and suggest strategies to address community-level barriers to HIV engagement along the care continuum.

## 5 CONCLUSIONS

The findings of our study revealed significant spatial heterogeneity in the HIV care continuum in Zambia. Despite similar individual-level factors associated with each stage of the continuum, our results demonstrated distinct geographic patterns for each “95,” which were not necessarily linked to high HIV prevalence areas. Our study underscores the need for targeted interventions to improve HIV status awareness, ART uptake, and viral load suppression in vulnerable populations. Although HIV diagnosis represents the most significant gap in the care continuum, a tailored approach may be required in different subnational regions to achieve the “95-95-95” targets. Further investigation into the drivers of care continuum gaps, including community vulnerability, can inform the development of innovative strategies to enhance HIV care continuum outcomes. Such strategies should take into account the unique challenges faced by different regions and populations, such as rural residence, low income, and limited healthcare access, to improve the effectiveness of HIV care and treatment programs. Addressing these gaps in the HIV care continuum is critical for reducing the burden of HIV and achieving global HIV targets to ensure that no one is left behind.

## Notes

### Competing Interest Statement

The authors have declared no competing interest.

### Author Declarations

Data are available in a public, open-access repository. The data that support the findings of this study are available from the Population-Based HIV Impact Assessment (PHIA; https://phia-data.icap.columbia.edu/). Human subjects and ethical approval for the ZAMPHIA survey was granted by the Zambia National Health Research Ethics Board, the Tropical Diseases Research Centers Ethics Review Committee, and the Institutional Review Boards at the Centers for Disease Control and Prevention (CDC; Atlanta, Georgia, USA). All participants provided written informed consent or assent for the original survey.

